# Factors affecting use of speech testing in adult audiology

**DOI:** 10.1101/2021.10.15.21265052

**Authors:** Bhavisha J Parmar, Saima L Rajasingam, Jennifer K Bizley, Deborah A Vickers

**Author notes:** UCL Ear Institute, University College London, 332 Grays Inn Road, London, WC1X 8EE.

## Abstract

**Objective:** To investigate the factors affecting the use speech testing in adult audiology services

**Design:** A mixed-methods cross-sectional questionnaire study

**Study Sample:** A UK sample (*n*=306) of hearing healthcare professionals (HHPs) from the public (64%) and private sector (36%) completed the survey

**Results:** In the UK, speech testing practice varied significantly between health sector. Speech testing was carried out during the audiology assessment by 68% of private sector HHPs and 5% of those from the public sector. During the hearing aid intervention stage speech testing was carried out by 40% and 8% of HHPs from the private and public sector, respectively. Recognised benefits of speech testing included: 1) providing patients with relatable assessment information, 2) guiding hearing aid fitting, 3) supporting a diagnostic test battery. A lack of clinical time was a key barrier to uptake.

**Conclusion:** Use of speech testing varies in adult audiology. Results from the present study found the percentage of UK HHPs making use of speech tests was low compared to other countries. HHPs recognised different benefits of speech testing in audiology practice but the barriers limiting uptake were often driven by factors derived from decision makers rather than clinical rationale. Privately funded HHPs used speech tests more frequently than those working in the public sector where time and resources are under greater pressure and governed by guidance that does not include a recommendation for speech testing. Therefore, the inclusion of speech testing in national clinical guidelines could increase the consistency of use.

## Introduction

Difficulties understanding speech in the presence of noise or competing speakers is one of the most common complaints of people with hearing loss and hearing aids (Abrahms, 2015). Never-the-less, the main assessment of hearing sensitivity is pure tone audiometry. Pure tone audiometry does not always effectively predict speech perception (De Sousa et al., 2020; Liberman, 2017; Vinay & Moore, 2007) because it indicates a listener’s access to sound rather than their functional hearing ability. The discrepancy between common clinical practice and patient-reported priorities can result in lower patient satisfaction or poor hearing aid usage.

Within audiology practice, hearing healthcare professionals (HHPs) may choose to perform speech tests for a variety of reasons, depending on clinical requirements. They are also commonly used as an outcome measure in auditory research studies e.g. investigating benefits of hearing devices (Bosen et al., 2021; Ricketts & Picou, 2021) or effects of auditory training (Burk et al., 2006; Zhang et al., 2021). Speech tests include the measurement of an individual’s speech recognition thresholds and responses to supra-threshold speech in aided and/or unaided testing conditions, in quiet or in noise. Such methods can be beneficial when choosing amplification devices, setting patient expectations and providing onward referral e.g. cochlear implant candidacy (Turton et al., 2020). They are also recommended for use prior to hearing aid fitting to capture a listener’s functional ability and identify appropriate intervention strategies (Ricketts, 2019). Assessing speech perception abilities in the presence of noise may better reflect the listening conditions that patients report as more challenging (Carhart & Tillman, 1970; Smits & Houtgast, 2005) and a range of commercially available speech-in-noise (SIN) tests are available to help to quantify abilities (e.g., QuickSIN (Killion et al., 2004), Bamford-Kowal-Bench (BKB) SIN (Bench et al., 1979; Niquette et al., 2003), HINT (Hearing in Noise Test) (Nilsson et al., 1994). A recent systematic review, evaluating behavioural assessment methods used before hearing device fitting, reported that patients who underwent SIN testing were more likely to have higher measures of hearing aid satisfaction (Davidson et al., 2021). According to a global survey of audiology practice, audiologists in 46% of countries (n = 62 countries, representing 78% of the world’s population) carried out speech tests (Goulios & Patuzzi, 2008) (See Supplemental Materials Table 1 for a summary of international speech test use). Speech testing is also used within cochlear implant (CI) candidacy assessment in the UK, but such practice in other countries varies. This may be driven by the differing service delivery models and funding sources for CI assessment and rehabilitation as well as a lack of clear clinical guidance in many countries (British Cochlear Implant Group, 2017; Vickers et al., 2016a). The inconsistency of practice is particularly concerning as preoperative level of speech understanding is one of the most valuable measures within the CI referral and candidacy assessment (Vickers et al., 2016b; Zwolan et al., 2020).

Audiology-related healthcare financing and delivery vary globally; while most patients worldwide rely on private insurance or self-funded care, many countries (e.g., UK, Australia, Norway, Sweden, Belgium, France) provide public insurance for audiology services (Moller, 2016; Yong et al., 2019). HHPs’ training and education also differs significantly across the world (Goulios & Patuzzi, 2008), and between private and public sectors, which can impact consistency and comparison of service provision. For instance, an Audiologist in the USA qualifies with a Doctorate in Audiology (AuD) whereas a hearing aid dispenser in the UK, can register with the Health and Care Professions Council (HCPC) with a foundation degree (FdSc) and internship (or an equivalent qualification). Currently, a public sector Audiologist, working in the National Health Service (NHS) in the UK, will require an undergraduate degree in audiology, or its equivalent. Some countries include speech testing within the scope of recommended audiology practice (College of Audiologists and Speech-Pathologists of Ontario, 2018; Rehabilitation Council of India, 2015), whereas others do not (British Academy of Audiology, 2014). Health policies for England and Wales are based upon guidance produced by the National Institute for Health and Care Excellence (NICE) and such guidance may also have a wide influence on the development and implementation of global clinical practices (Chandra et al., 2015; van der Straaten et al., 2021; Vasse et al., 2012; Yue et al., 2014). The latest NICE guidance for the assessment of adults with hearing difficulties does not include any recommendations for speech testing with this population (National Institute for Health and Care Excellence, 2018). Such guidance results in relevant resource allocation being cut for speech testing leading to individual services deciding on whether they can accommodate speech testing in their audiology provision. This can result in discrepancies across service delivery in audiology practice. Further evidence is required to evaluate if there is a role for speech testing in audiological practice, and what form that it should take.

The aim of this research was to evaluate hearing healthcare professionals’ (HHPs) speech testing practices in routine adult audiology services within the UK, and better understand the facilitators and barriers to speech testing provision. This work is presented within the framework of UK audiology healthcare delivery for both public and private practice. This approach enables comparison with other countries based on a public or private funding infrastructure.

## Methods

### Ethical approval

This study was approved by the University College London Ethics Committee (Project no. 3866/001). This research also received internal ethical approval from all professional organizations that assisted in questionnaire distribution. All questionnaires were completed anonymously, and respondents were not asked to provide any personally identifiable details or health information. Data was stored in compliance with the European Union’s General Data Protection Regulation (2016/679) and participant consent was implied based on their completion of the questionnaire.

### Recruitment

HHPs who were providing audiological care to adult patients in the UK were invited to take part in the online questionnaire. The study information sheet and questionnaire hyperlink were circulated to the audiology departments of all public sector hospitals as well as members of British audiology societies and professional bodies. HHPs were also asked to forward the questionnaire on to colleagues or other HHPs they knew were working in the audiology field. The questionnaire remained open to respondents for 12 weeks between April and June 2019.

### Questionnaire Development

A questionnaire was designed specifically for this research study, customized to address three main areas of interest for the responding HHPs: 1) *demographics* (i.e., employment sector, main patient population, and geographical location), 2) *speech testing* (i.e., the equipment, frequency, and type of speech testing carried out at a patient’s initial assessment and hearing aid intervention), and 3) *Opinions of using speech testing in audiology* (i.e., benefits and barriers to performing speech tests). Given that longer surveys are less likely to be completed (Sahlqvist et al., 2011), the present questionnaire was designed to ensure that all questions could be accurately completed within five minutes. During the pilot stage, four expert HHPs reviewed the clarity and content of the questionnaire and modifications were made based on these comments.

The final version of the questionnaire contained eleven questions (see Supplementary Materials): ten multiple choice questions and one open-ended question (“What are the benefits of speech testing in adult audiology?”) inviting a free text response. The survey was developed, distributed, and completed via the SurveyMonkey web-based tool and all anonymized data were securely stored. Although respondents were able to skip questions they preferred not to answer, only 21 respondents utilised this function and all results are presented alongside the total number of respondents for each question.

### Response Rate

Overall, 306 HHP respondents completed the online questionnaire. Eleven respondents reported only working in paediatric audiology or academia and were therefore removed from the analyses. This resulted in a total of 295 HHPs providing valid questionnaire data for the study. This sample size reflects approximately 8-10% of UK registered HHPs and is similar to the number of respondents obtained by other UK-based surveys of audiologists (Parmar et al., 2021; Wright et al., 2014).

### Data analysis

As HHPs may have received multiple invitations to participate in the present research study, IP addresses were checked to ensure respondents only completed the questionnaire once. Descriptive statistics were used to summarize the multiple-choice questionnaire items. Chi-squared (χ^2^) tests were used to compare speech testing occurrence across sectors (public vs. private). For Chi-squared statistical analysis, the Likert-scale responses “Often” and “Always” were combined to present the proportion of HHPs regularly conducting the activity and categories “Never” and “Rarely” were combined to present those that rarely conducted the activity. Thematic analysis was used to analyse free text responses (to the open-ended question) by grouping statements into codes and themes (Braun & Clarke, 2006). Inductive thematic analysis allows for the extraction of emerging codes from the data throughout the analysis process (i.e., data-driven approach), rather than relying on a pre-determined list of themes that are used to interpret the data.

## Results

### Demographics

Responses from 295 HHPs actively practicing within adult audiology services, across the UK, are shown. There were 64% of respondents working in the public sector (National Health Service) and 36% working in the private sector.

### Speech tests in routine adult audiology

Overall, 26% of HHPs reported carrying out speech testing at the first audiological assessment appointment (see Figure 1) but only 18% of respondents indicated that they “often” or “always” carry out speech testing during hearing aid fittings and follow-up appointments.

**Figure 1.**
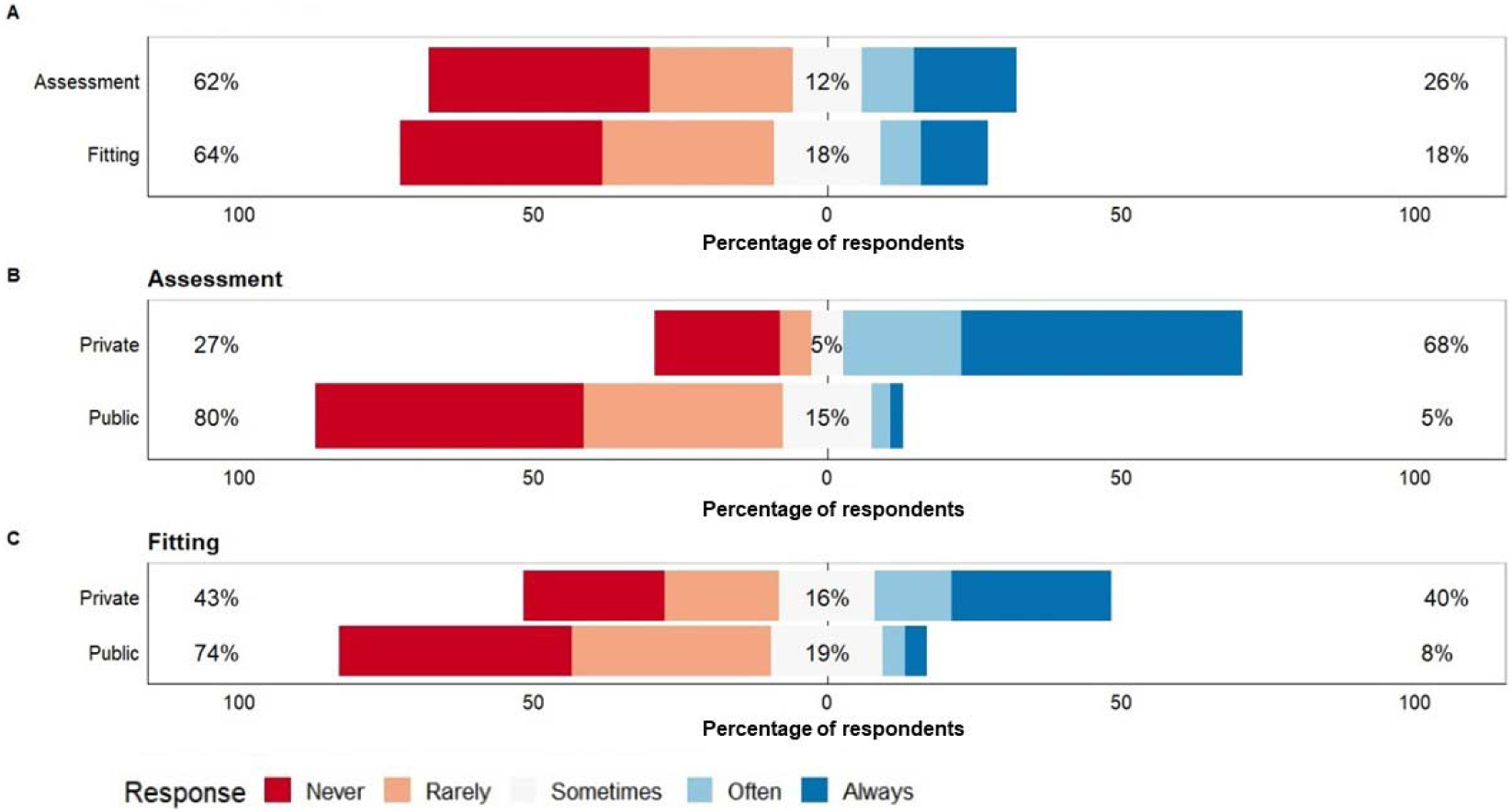
The use of speech testing in adult audiology (question 3&5 of the survey). (A) The distribution of HHP respondents’ use of speech testing during adult audiology assessments and fittings, (B) Private and public sector HHPs’ use of speech testing during the hearing assessment & (C) The use of speech testing during hearing aid fittings by private and public sector HHPs.

### Speech testing in private vs. public sector practices

As evident in Figure 1, there was a significant effect of sector type on speech testing practice (χ^2^ (1, *N* = 247) = 116.68, *p* < 0.001), with HHPs from the private sector using speech testing more often than HHPs from the public sector. This was present at both the initial assessment stage (private sector: 68% vs. public sector: 5%) as well as during hearing aid intervention (private sector: 40% vs. public sector: 8%).

### Types of speech tests used in adult audiology practice

The QuickSIN and AB word lists were the most commonly used speech test materials for the initial assessment appointment and the hearing aid intervention (Table 1). Four alternative speech measures were identified by respondents as “Other” these were: the City University of New York sentences test (Boothroyd, 1985), the Acceptable Noise Level test (Nabelek et al., 1991), the Ling sounds (Ling, 1976)) and informal conversational speech with and without lip reading cues.

**Table 1.**
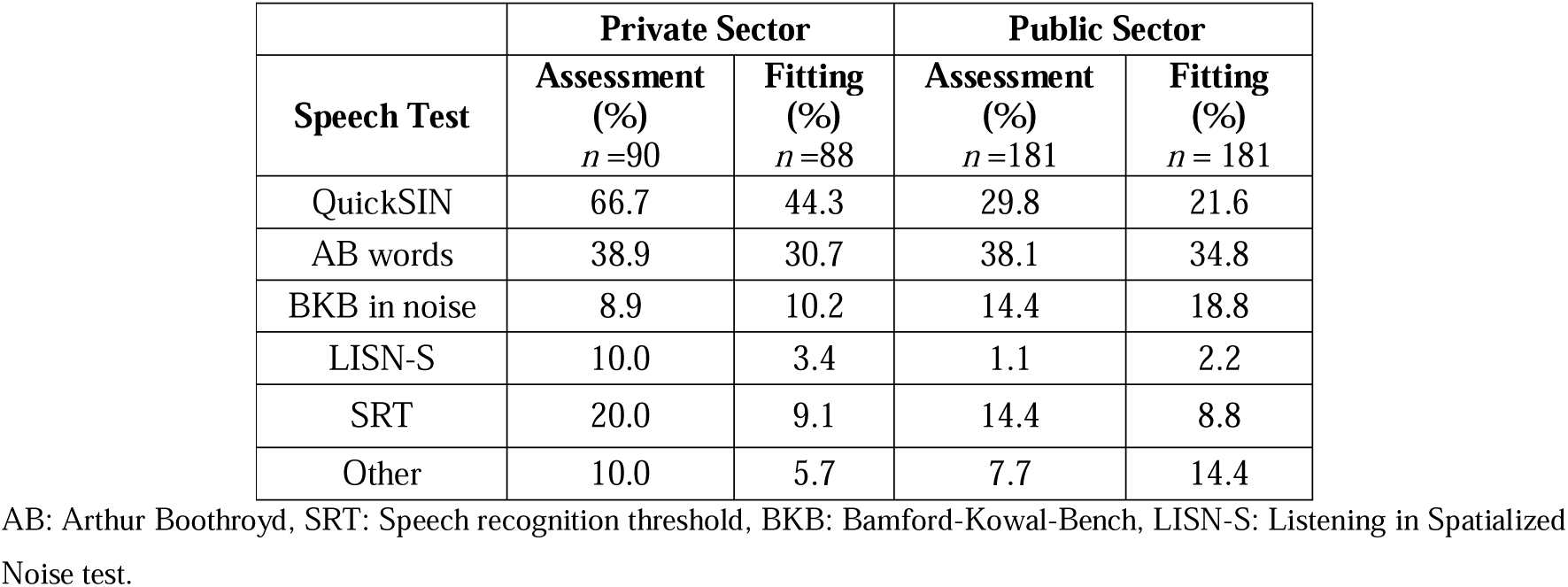
Speech test materials used at the first audiological assessment appointment and/or follow-up appointments (% of responses).

| Speech Test | Private Sector |  | Public Sector |  |
| --- | --- | --- | --- | --- |
|  | Assessment (%)<br>*n* =90 | Fitting (%)<br>*n* =88 | Assessment (%)<br>*n* =181 | Fitting (%)<br>*n* = 181 |
| QuickSIN | 66.7 | 44.3 | 29.8 | 21.6 |
| AB words | 38.9 | 30.7 | 38.1 | 34.8 |
| BKB in noise | 8.9 | 10.2 | 14.4 | 18.8 |
| LISN-S | 10.0 | 3.4 | 1.1 | 2.2 |
| SRT | 20.0 | 9.1 | 14.4 | 8.8 |
| Other | 10.0 | 5.7 | 7.7 | 14.4 |

AB: Arthur Boothroyd, SRT: Speech recognition threshold, BKB: Bamford-Kowal-Bench, LISN-S: Listening in Spatialized Noise test.

Ear-specific transducers (either insert or supra-aural headphones) were the most commonly used equipment for speech testing (see Table 2). The use of one individual loudspeaker (39.7%) or a live voice in a face-to-face context (24.4%) were also common. Three alternative methods (live voice of the patient’s family member, live voice with a sound level measurement, and bone conduction) were identified as “Other”.

**Table 2.** Equipment used during speech testing in adult audiology (% of responses).

| Equipment Type | Public sector (%)<br>(*n* =188) | Private sector (%)<br>(*n* = 94) | Total (%)<br>(*n* = 282) |
| --- | --- | --- | --- |
| **Ear specific transducers (Headphones/inserts)** | 55.3 | 44.7 | 51.7 |
| **Live voice (face-to-face)** | 25.0 | 23.4 | 24.4 |
| **1 loudspeaker** | 39.8 | 42.2 | 39.7 |
| **2 loudspeakers** | 10.2 | 7.8 | 9.2 |
| **Multi speaker array** | 2.2 | 6.7 | 3.5 |
| **Other** | 7.1 | 8.7% | 4.3 |

### Barriers to using speech testing in routine adult audiology

The most commonly reported barrier to performing speech testing was the lack of clinical time (59.8% of all respondents). This was followed by the lack of appropriate equipment and a lack of training (Table 3). In addition to the five forced-choice barrier options offered in the questionnaire (lack of clinical time, lack of equipment, lack of training, lack of test sensitivity and lack of benefit), HHPs identified several other factors that they felt posed significant barriers to speech test use. These included the lack of speech test material in non-English language, unfamiliar accents, departmental protocol restrictions, a lack of normative data, a limited availability of tests suitable for adults with additional needs, and no clear evidence for patient benefit or care.

**Table 3.** Barriers to completing speech testing in adult audiology.

| **Barriers** | **Public Sector**<br>**(%)**<br>***n* = 188** | **Private Sector**<br>**(%)**<br>***n* = 90** | **Total**<br>**(%)**<br>***n* = 291** |
| --- | --- | --- | --- |
| **Lack of clinical time** | 78.1 | 24.4 | 59.8 |
| **Lack of appropriate equipment** | 34.6 | 16.7 | 28.1 |
| **Lack of training** | 21.3 | 8.9 | 16.5 |
| **Lack of test sensitivity** | 6.4 | 10 | 7.6 |
| **Lack of benefit** | 5.3 | 1.1 | 3.8 |
| **Other** | 12.8 | 16.7 | 13.8 |

### Benefits of speech testing

Responses to the open question “What do you think the benefits of using speech testing in routine adult audiology are?” were analysed thematically. Although this question asks for benefits of speech testing, and therefore indicates the need for positive responses, some respondents specified ‘N/A’ in the response area if could not name any benefits (See Supplemental Materials for all responses). Three key themes were identified: 1) *providing patients with relatable assessment information*, 2) *guiding hearing aid fitting* and 3) *supporting a diagnostic test battery*. All themes were present in responses by HHPs from both the public and private sectors. A high number of respondents in both the public and private sector HHP groups indicated that speech tests provided an important tool for helping to demonstrate and explain audiometric and speech perception in relation to everyday listening difficulties (52% of public sector HHPs & 53% of private sector HHPs) and also for guiding hearing aid fitting (41.4% of public sector & 40% of private sector). Private sector practitioners less commonly suggested that speech testing was helpful in diagnostic test batteries compared to public sector respondents (27.3% of public sector vs. 16.7% of private sector).

#### 1) Providing patients with relatable assessment information

Speech testing provides patients and their families with information about the personal impact of hearing loss on speech understanding. The respondents thought that speech testing was particularly helpful for demonstrating functional hearing difficulties to patients (and their families) and provides the HHP with another tool to identify patients’ communication barriers.

> *“Convinces more than any other test that the patient’s problem is: a) real; b) serious. Denial is the biggest barrier to acceptance - speech testing breaks the barrier down better than everything else combined!”* (Private sector)
>
> *“For patients, it highlights need and reaffirms their awareness of hearing loss. For clinicians, it helps us understand their communication issues in a way a pure tone audiogram cannot*.*”* (Private sector)

HHPs reported that presenting speech to patients with hearing loss was more effective than pure tone audiometry alone for helping patients relate testing results to real world listening scenarios.

> *“Real life stimulus is often easier for the patient to relate to, and it also gives a better idea of the actual benefit to the patient. Also gives some idea of processing as opposed to just detection of sound*.*”* (Public sector)
>
> *“People will often associate the test with their primary issues with background noise and will often feel listened to when the speech test is complete and explained*.*”* (Private sector)
>
> *“The standardised audiogram doesn’t reflect realistic listening situations and so speech tests may provide a more accurate representation of an individual’s hearing difficulties”* (Public sector)

Finally, respondents felt that speech testing helped HHPs manage patients’ expectations during audiology consultations. One commonly reported example was the mismatch in the audiogram classifications (e.g., mild hearing loss) and the communication difficulties experienced by the patient. Therefore, they believed that speech testing was beneficial for improving patients’ understanding of their own hearing abilities. Beyond diagnostic assessment, speech testing was also reported to be beneficial for the counselling process and could lead to improved patient satisfaction.

> *“It allows you to gain a picture of their actual hearing. A person can have a mild loss but may struggle with speech more so. It is also a good tool to use for counselling and rehab purposes and allows you to set realistic expectations*.*”* (Public sector)
>
> *“Gives you a better understanding of patients loss and possible problems, for both clinician, patient and family which should allow for better treatment outcome. Should reduce return appts for some patients and lead to better satisfaction for those often listed as difficult*.*”* (Public sector)

#### 2) Guiding hearing aid fitting

Speech tests can be used to measure functional hearing aid benefit. HHPs reported making hearing aid adjustments based on speech tests and the results from aided speech testing were used to identify the limitations of hearing aids. Although HHPs felt speech testing was a useful counselling tool to demonstrate aided vs unaided performance, they reported a lack of standardisation in assessing hearing aid benefit.

> *“Gives level of functional hearing difficulty perhaps not revealed by PTA alone. Allows before and after aiding comparison to validate efficacy of hearing aids*.*”* (Public sector)
>
> *“A really useful counselling tool with patients at assessment to explain their processing in noise without visual cues (especially those with a greater SNR loss). A shame it is not a verified test for assessing improvement with hearing aids*.*”* (Private sector)
>
> *“Speech testing is a good tool to measure the benefit from the hearing aid. If speech testing is carried out on the first fit appointment, we can adjust the hearing aid characteristics based on this and address some issues then and there which will improve patient satisfaction and can reduce follow up visits*.*”* (Public sector)
>
> *“It’s the only way to understand the functional impact. It can be very surprising how much the speech test scores can vary from the audiogram in patients who are not successful with amplification*.*”* (Private sector)

#### 3) Supporting a diagnostic test battery

Speech testing was also reported to be a useful part of the audiological test battery to assist in the diagnosis of specific auditory conditions, especially in those where the patient’s functional hearing is not predictable from the audiogram (e.g., auditory processing disorder, non-organic hearing loss).

> *“Diagnostics, part of APD (*Auditory Processing Disorder*) test battery, aural rehabilitation (cognitive load and Listening effort, directional microphones, noise reduction algorithms, validation of fitting etc*.*)”* (Private sector)
>
> *“We are sometimes asked by ENT to perform speech testing on acoustic neuroma clinics, but do not routinely use them unless there is a complex patient or unexplained problems with hearing, i*.*e*., *to assess if there may be an auditory processing issue”* (Public sector)

HHPs also reported the use of speech testing in the audiological diagnostic assessment battery for determining cochlear implant candidacy (indications for cochlear implants).

> *“Identifies patients with very poor speech discrimination abilities. Assists with counselling on cochlear implant candidacy”* (Public sector)

## Discussion

The main aim of this research was to understand the factors affecting the provision of speech testing practice in UK adult audiology, within the private and public sector. By separating our sample into public and private sectors we were able to highlight how these are influenced by different factors. In general, the private healthcare sector is consumer-oriented and quality services are underpinned with the understanding that the consumer can withhold resources at their discretion, which can have significant implications to the future development and functioning of the organisation (Herrera et al., 2014). The public health sector in the UK (National Health Service: NHS), however, is clinician/systems-centred and services are driven by professional protocol and national clinical guidance rather than end-user review (Bradshaw & Bradshaw, 2004; Shen et al., 2007). In recent years, however, European public healthcare systems have adapted to increase the choice of healthcare provider available to the patient, with the assumption that a competitive market would improve the overall quality of services (Walumbe et al., 2016). In England, the ‘Any Qualified Provider’ policy was established to allow a specific subset of patients to choose any audiology provider (NHS services, private sector or voluntary sector), as long as they met an agreed quality standard and price (Department of Health, 2011). Given the continuously adapting nature of healthcare service delivery models and national clinical guidance, it is important to explore factors that influence audiological clinical practice across sector, including the use of speech testing. This is particularly important as private hearing aid services for adult patients in the UK, are steadily growing (The British Irish Hearing Instrument Manufacturers Association, 2021).

Overall, our results for the rate of use of speech tests in UK adult audiology practice (26% during assessment; 18% during hearing aid fitting), are far lower than indicated in the literature on audiology practice data from the USA (American Speech-Language-Hearing Association, 2019; Kirkwood, 2005), Canada (DeBow & Green, 2000), Australia (Myles, 2017), South Africa (Thakor, 2020), and Saudi Arabia (Alanazi, 2017) (see Supplemental materials Table 1). However, these results mask a significant difference in speech testing approaches between the public and private sector, with more HPPs in private sector being more likely to perform speech testing than those in the public sector. The type of test used also differed with sector and appointment type. This variability is evident around the world. For instance, SIN administration rates in India vary from 4.5% of respondents (Easwar et al., 2013) to 34% (Nandurkar et al., 2015) and SIN measures were not performed by any respondents in the Saudi Arabian study (Alanazi, 2017). In contrast, a study of American audiologists found that 66% incorporate aided SIN measures and 80% performed unaided SIN measures at the initial hearing aid fitting (Anderson et al., 2018). Differences in healthcare provision models across the world, may partially explain these discrepancies. More specifically, audiology practices within the USA are most akin to the reports from private sector UK-based HHPs, given that American health services are predominantly reliant on private funding and insurance policies. These findings are further supported by a study demonstrating speech testing as a treatment outcome after hearing aid fitting in private American audiology practices (American Speech-Language-Hearing Association, 2019). It can be argued that the commercial nature of private sector hearing aid provision may influence the provision of speech testing.

Despite the low uptake of speech tests in the UK, respondents reported specific speech test measures and equipment used in clinical practice. Ear-specific transducers were the most commonly used equipment used for delivering speech testing, although in many cases a single loudspeaker set-up was also used. Surprisingly, reliance on face-to-face live voice was identified as the third most popular choice for speech testing modality. The prevalence of this method was unexpected given the uncalibrated, highly variable nature of interactive, naturalistic voice testing which can lead to inconsistencies between HHPs and make it very challenging to compare performance between testing conditions and testing centres (Hood & Poole, 1980; Roeser, 2008). Audiologists in Canada (DeBow & Green, 2000), South Africa (Thakor, 2020) and Australia (Myles, 2017) also reported a high reliance on the use of live voice during speech testing. The increased use of live voice testing may be due to variation in equipment, lack of validated recorded speech materials in appropriate languages/accents and time availability.

Audiology surveys carried out in Australia, South Africa and India highlighted the reasons why HHPs carried out speech testing in routine adult audiology practice (Myles, 2017; Nandurkar et al., 2015; Thakor, 2020). These included: cross-checking results with pure tone audiometry findings, counselling and managing patient expectations, assessing hearing aid candidacy and use within the diagnostic test battery. In the present study, despite the relatively low uptake of speech testing across adult audiology practices in the UK, HHPs reported different beliefs in the potential benefits of speech testing. One of the most common benefits was the way speech testing helped patients and their families understand audiological assessment results in relation to their everyday speech perception difficulties. Adults with hearing loss have previously reported that HHPs were not in tune with their communication needs and patients could not recall technical clinical information (Watermeyer et al., 2015). Using a more ecologically valid stimuli, like speech, could help patients apply their diagnostic results to their real-world listening scenarios. Previous literature has suggested that enhanced ecological validity can lead towards more integrated and individualised hearing healthcare (Keidser et al., 2020).

Several HHPs indicated that speech testing was beneficial to compare functional performance pre- and post-hearing aid fitting. This included the ability to validate the efficacy of the hearing aid fitting, and to adjust the hearing aids based on the hearing aid settings. However, some respondents reported the lack of verified methods to measure hearing aid benefit. Previous studies have used speech testing as a sensitive outcome measure to explore the impact of complex hearing aid systems (Glista et al., 2009; Wolfe et al., 2011), and to evaluate hearing device fine tuning (Tonelini et al., 2016). However, current speech testing practice guidance does not include the adjustment of hearing aids in response to speech testing results (British Society of Audiology (BSA), 2019). Clinical HHPs may benefit from further training and guidance to meaningfully interpret and use unaided and aided speech testing results to assess and improve the hearing aid fitting and rehabilitation.

Respondents, particularly those from the public sector, reported the importance of using speech testing within the clinical diagnostic test battery. This could be due to the public sector’s connection to other medical departments e.g., tertiary level audiology department being connected to Ear, Nose and Throat and cochlear implant centres. In such settings there may be additional need for speech testing. The differences in HHP training and education between private and public sector in the UK may also contribute for differences in identifying need for speech testing based on medical rationale. Speech testing is included in the diagnostic test battery for specific conditions e.g., central auditory processing disorder (American Speech-Language-Hearing Association, 2014). Therefore, it is important for HHPs, regardless of sector, to remain up to date with information about their use and have access to adequate training and resources to provide appropriate intervention options for patients. Furthermore, as speech testing is performed within the cochlear implant candidacy assessment across the world (British Cochlear Implant Group, 2017). Raising awareness of speech testing within routine audiology practice could help identify potential candidates earlier and lead to increased uptake.

Despite the overall low uptake of speech testing presented in the current study, the lack of benefit of such testing was not considered a common limiting factor. The majority of public sector HHPs in the UK reported the lack of clinical time as a key barrier to performing speech testing, despite the availability of assessments that are designed to be completed within a few minutes (e.g. QuickSIN). In the UK, public health commissioning groups use NICE guidelines to allocate funds and resources (Chundu & Flynn, 2014) and the absence of speech testing in such guidelines could influence the time and resources allocated for these activities in public sector audiology services. Audiologists around the world report a lack of government funding for audiology services (Goulios & Patuzzi, 2008). Previous research has also found time demands to be the highest stress factor for HHPs (Emanuel, 2021; Severn et al., 2012), but some suggest these factors may affect more public sector clinicians than independent private clinicians (Mott et al., 2004). The flexible resource management of the private sector is likely to impact the significant differences between private and public sector speech testing provision observed in this study and globally.

Beyond the multiple-choice list of barriers, respondents also reported several additional factors limiting speech testing uptake, including the absence of speech testing in departmental protocols and a lack of evidence for how speech testing can be used for individual patients. These align with barriers reported worldwide (See Supplemental Materials Table 1). Although speech testing is recognised as a functional hearing assessment, the lack of standardization poses a barrier to its clinical use (Moore et al., 2019). This will affect the consistency of speech testing usage between clinicians and services across the world. The use of speech materials are sensitive to a person’s cognitive function (Nuesse et al., 2018) and the choice of using word or sentence stimuli is dependent on clinical requirement and the influence of other factors e.g. contextual cues in sentence materials (Wilson & McArdle, 2005). Speech tests differ in functionality in a hierarchical manner; some assess the listeners’ ability to detect speech stimuli whereas others assess sentence discrimination. Therefore, HHPs require guidance to choose the most appropriate measure, depending on evidence and clinical need. There is also need for more sensitive triage techniques and cognitive screening measures within audiology to improve the holistic interpretation of results (Shen et al., 2016). Speech test materials are not available for all languages and accents (Nandurkar et al., 2015; Thakor, 2020). These barriers were also reported by UK respondents to the present study. However, there are recommendations for the construction of multilingual speech tests available so that the test can be carried out in the listener’s native language even if the tester does not understand that language (Akeroyd et al., 2015). The absence of speech testing in departmental and national guidance was reported by HHPs in the present study, in agreement with findings from other countries (Alanazi, 2017). The lack of such guidance may contribute to inconsistency in practice, including the use of speech testing. A collaboration between health authorities, researchers, hearing device manufacturers, HHPs and service users could result in the development of accessible toolkits of validated speech test materials, normative data, and recommended equipment as well as practical guidance.

### Strengths, limitations, and future directions

This research is the first of its kind to report on the patterns of speech testing practice in routine adult audiology practices, within the UK and considering the impact with an international perspective. A major strength of this work is the inclusion of both public sector and private sector audiology services and the placement of UK-based within an international context to unveil the similarities and differences in audiology practices around the world. The provision of healthcare services by private versus public sectors differs country-to-country, necessitating a direct comparison of these different service delivery approaches on an international scale. There are also some limitations within the current study. Due to the sampling method, it is not possible to calculate a response rate for the current study. Furthermore, although many UK-based HHPs felt speech testing was beneficial in the present study, this research did not investigate the reasons and motivations for using speech testing on a case-by-case basis and whether the incorporation of speech testing impacts patient care, outcomes, and/or satisfaction, as well as hearing aid use. Data exploring clinical audiology service provision is often collected by professional bodies or incorporated within grey literature (See Supplemental Materials Table 1). It would be beneficial for future work to include the distribution of an international questionnaire of audiology practice, including the use of speech testing, in collaboration with professional bodies and HHPs for dissemination through peer reviewed publication. The data collection for the current study was carried out before the COVID-19 pandemic but current audiology service provision will reflect the increased use of remote care/teleaudiology, and therefore a reduction in speech testing. Future work could include the exploration of how audiology services have developed in response to the pandemic and the remote care options available for speech testing and the use and uptake of such testing.

## Conclusion

While pure tone audiometry gives information about a listener’s hearing sensitivity, HHPs report speech testing to be beneficial in providing patients with relatable information about their functional hearing, to guide the hearing aid fitting and to use within the diagnostic test battery. The present research demonstrates that the global provision of speech testing is variable, with the UK demonstrating relatively infrequent use of speech testing during the clinical assessment and hearing aid fittings in adult patients by public sector HHPs. Private sector audiology practices in the UK, however, were more comparable to uptake reported for the USA and Canada. A lack of clinical time, training and equipment were identified as primary reasons affecting provision variability in the UK and likely to also account for global heterogeneity in service provision. Given the evolution of new audiological assessment techniques, it is important to gather data of current clinical practice trends. Clinical practice guidance could be developed to enhance consistency of speech testing methods and recommend relevant training and resources for HHPs around the world. The inclusion of speech testing within the formal scope of practice for audiologists and within clinical practice guidance could facilitate the allocation of necessary resources for public sector HHPs in the UK and beyond.

## Data Availability

All data produced in the present study are available upon reasonable request to the authors

## Acknowledgements

We would like to thank the British Society of Audiology, British Academy of Audiology, and the British Society of Hearing Aid Audiologists for distributing this survey. Our thanks are extended to the respondents who completed the survey.

