## Supplemental information for "Factors affecting use of speech testing in adult audiology"

**Table 1.** A summary of survey study results provided by HHPs in relation to the implementation of speech testing practices in adult audiology across the world.

| **Study** | **Country** | **HHPs** | **Use of Speech Tests** | **Transducer** | **Use/Barriers** |
| --- | --- | --- | --- | --- | --- |
| (Martin et al., 1998) | USA | 218 | *SDT: 69%*  *SRT: 99.5%* | *Monitored live voice*: 94% | N/A |
| (DeBow & Green, 2000) | Canada | 115 | *Word recognition threshold measures: 85%* | *Monitored live voice:* 89%  *Supra-aural headphones:* 90% | N/A |
| (Kirkwood, 2005) | USA | 674 | Speech audiometry:  *Never: 1.2%, half the time: 1.2%, always: 90.8%* | N/A | N/A |
| (Easwar et al., 2013) | India | 199 | *SRT only: 24%*  *SRT & speech identification: 38.7%*  *SRT & SIN: 2.5%*  *SIN only: 2%*  *No routine speech tests:19%* | N/A | N/A |
| (Nandurkar et al., 2015) | India | 59 | Speech perception tests:  *Always: 22%, often: 34%, sometimes: 36%, rarely/never: 8%*  SIN: *Always: 5%, often: 29%, sometimes: 34%, rarely: 17%* | *Headphones:* 21%  *Sound-field:* 15% | Reasons for using speech tests: *assess hearing aid efficacy: 76%, hearing aid candidacy assessment: 63%, assessing patient difficulties: 52%, diagnostics: 39%*  Barriers: *Time constraints, lack of adequate material, lack of ideal setting, lack of proficiency in the client’s first language* |
| (Alanazi, 2017) | Saudi Arabia | 23 | SRT: *65%*  SDT: *48%*  SIN*: 0%* | N/A | N/A |
| (Ali et al., 2017) | Malaysia | 111 | Speech audiometry:  *Never: 62.24%, half the time: 26.53%, usually/always: 11.22%* | N/A | N/A |
| (Myles, 2017) | Australia | 312 | AB word lists:  *Routine use: 95%*  *In quiet: 99.6%, in noise: 5%* | Live voice: 2%, ear specific transducer: 66%, | Reasons for use:  *Cross-check pure tone audiogram:96%, diagnostic: 83%, counselling: 87%, protocol requirement: 63%, rehabilitative: 79%* |
| (American Speech-Language-Hearing Association, 2019) | USA | 751 | Implementation of SIN testing to validate treatment outcomes:  *Daily/weekly: 35%*  *Monthly: 26%*  *Never: 39%* | N/A | N/A |
| (Anderson et al., 2018) | USA | 251 | Initial hearing aid fitting:  *SRT & word recognition: 98%*  *Unaided SIN: 80%*  *Aided SIN: 66% (often or sometimes)*  Fine tuning of hearing aids:  SIN*: 67%*  Speech-in-quiet*: 66% (often or sometimes)* | N/A | N/A |
| (Thakor, 2020) | South Africa | 107 | SRT: *Never: 13%, rarely: 7%, occasionally: 5%, sometimes: 7%, frequently: 9%, usually: 12%, always: 47%*  SIN: 36% | Live voice: 82%  Pre-recorded: 8% | Use of SRT: *calculating the correlation with the PTA, part of departmental/practice assessment protocol, to obtain a level from which to calculate the presentation level for other speech tests, counselling tool.*  Use of SIN: *patient counselling, managing patient expectations.*  Barriers: *language differences, lack of equipment, time constraints* |

SRT: Speech recognition threshold, SDT: Speech detection threshold, SIN: Speech-in-noise tests, AB: Arthur Boothroyd

**Survey: Speech testing in Adult Audiology (UK)**

1) Do you work in an NHS or private sector audiology department?
   **(This survey is only suitable for clinicians working in the U.K.)**
   a. Private sector
   b. NHS (public sector)
   c. Other (please specify)
2) What type of patients do you assess in your clinic?
   a. Adults
   b. Children
   c. Both adults and children
3) When seeing an adult patient in audiology for the first time do you perform any kind of speech perception testing as part of the assessment process?
   a. Never
   b. Rarely
   c. Sometimes
   d. Often
   e. Always
4) If applicable- What type of speech tests do you perform at the first (assessment) appointment?
   a. Speech audiometry (speech recognition threshold)
   b. Quick-SIN
   c. Lisn-S
   d. AB words
   e. BKB Speech in Noise Test
   f. N/A
   g. Other (please specify)
5) When fitting an adult patient with hearing aids do you perform any kind of speech testing on the day of the fitting or at the follow up appointment?
   a. Never
   b. Rarely
   c. Sometimes
   d. Often
   e. Always
6) If applicable- What type of speech testing do you carry out at the time of hearing aid fitting or follow up?
   a. Speech audiometry (speech recognition threshold)
   b. Quick-SIN
   c. Lisn-S
   d. AB words
   e. BKB Speech in Noise Test
   f. N/A
   g. Other (please specify)
7) If you perform speech testing, what do you use to present the speech tokens and noise?
   a. Live voice (your own voice rather than through speakers or headphones)
   b. 1 calibrated speaker
   c. 2 calibrated speakers to present speech and noise from separate locations
   d. Multi speaker array to present speech and noise from multiple locations around the patient
   e. Headphones/Inserts
   f. N/A
   g. Other (Please specify)
8) What are your main barriers/challenges to performing speech testing regularly?
   a. I do not have enough time in clinic
   b. I do not have the necessary equipment
   c. I do not see the benefit
   d. I do not have sufficient training
   e. The tests available are not sensitive enough to detect change (e.g. Changes in hearing aid features/programming)
   f. N/A
   g. Other (please specify)
9) What do you think the benefits of using speech testing in routine adult audiology are?

*Free text answer*

10) *Do you perform any tests of localisation for adult patients in your audiology clinic?* ***(this question was removed from analysis as publication focusses on speech testing only)***
    a. *Yes*
    b. *No*
    c. *If yes, please specify the types of tests you perform*
11) *Please select the geographical location of your workplace from the following options:*
    a. Scotland
    b. Wales
    c. Ireland
    d. England- Greater London
    e. England- South West
    f. England- South East
    g. England- Midlands
    h. England- North West
    i. England North East

| **Theme** | **Responses to question 9:**  **“What do you think the benefits of speech testing (for adult patients) are?”** |
| --- | --- |
| Providing patients with information | *It allows you to gain a picture of their actual hearing. A person can have a mild loss but may struggle with speech more so. It is also a good tool to use for counselling and rehab purposes and allows you to set realistic expectations.* [Public sector] |
|  | *Establish a realistic expectation of what benefit a hearing aid may provide .* [Public sector] |
|  | *Therapeutic benefit of ‘real life’ testing* [Private sector] |
|  | *Have an understanding of the patient's limitations before suggesting hearing aids, counselling*. [Private sector] |
|  | *Important for confirming handicap especially in noisy situations which is a very common with presbyacusis* [Private sector] |
|  | *Helps with counselling and setting the aids accurately.* [Public sector] |
|  | *Gives them support and better understanding* [Private sector] |
|  | *Proving loss and validity of aids* [Private sector] |
|  | *Useful habitation tool, counselling tool. Useful to demonstrate to user and family benefit. Useful tool to set realistic expectations and to work towards agreed goals to optimise hearing aid performance and work towards patient satisfaction* [Private sector] |
|  | *Counselling tool* [Private sector] |
|  | *Very important for them to understand the impact of their hearing loss and ultimately assist in motivation to correct* [Private sector] |
|  | *Great tool for counselling patients and family about expectations and nature of hearing loss and is more realistic, relatable to patients than pure tones.* [Public sector] |
|  | *Convinces more than any other test that the patient's problem is: a) real; b) serious. Denial is the biggest barrier to acceptance - speech testing breaks the barrier down better than everything else combined!* [Private sector] |
|  | *Real life is what is affecting them so why not use words as well as beeps.. also it's a great way to show them how context plays a main role in understanding people.* [Private sector] |
|  | *Great to prove hearing loss to px and support .* [Private sector] |
|  | *Better prediction of how well they will succeed with amplification Managing expectation Being aware of limitations of hearing aids .* [Private sector] |
|  | *Improved clinical decisions ..Can set realistic expectations* [Private sector] |
|  | *It helps show the client and the support that they are not hearing as well as they think they are.* [Private sector] |
|  | *I find that Quicksin is a really excellent tool for significant others to really appreciate the clients hearing loss and also I like to see the difference with aided and unaided results .* [Private sector] |
|  | *Being able to know the benefits of the hearing aids .* [Public sector] |
|  | *There are definitely benefits in terms of rehabilitation and helping patients take ownership of their problems (showing more objectively a before and after comparison).* [Private sector] |
|  | *Assessment of improvement in hearing ability in real world* [Private sector] |
|  | *It provides a qualitative measure of hearing ability which can reassure patients and also help family members understand the impact of a hearing loss on speech detection* [Public sector] |
|  | *Counselling tool if patients are struggling with discrimination* [Public sector] |
|  | *Shows benefits of aiding [Public* sector] |
|  | *It allows patients to physically see any improvements [Public* sector] |
|  | *People will often associate the test with their primary issues with background noise and will often feel listen to when the test is complete and explained.* [Private sector] |
|  | *Demonstrating benefit of hearing aids to patients. Subjective verification* [Private sector] |
|  | *To check the aids are giving the patient what they need in a more realistic way* [Public sector] |
|  | *Counselling, understanding benefits and limitations, monitor progression, awareness of deterioration in either hearing ability or function of aid/s.* [Public sector] |
|  | *provide valuable information about a person's hearing in noise for counselling* [Public sector] |
|  | *provides valuable information about a person's hearing ability.* [Public sector] |
|  | *Good for rehab for patients with processing issues and poor speech discrimination. Helps them to understand the issues.* [Public sector] |
|  | *Prediction of possible outcome of hearing intervention. Support for amount of benefit of hearing intervention . Change in performance using hearing intervention Help with further intervention or change in intervention decisions* [Public sector] |
|  | *Helps to show the patient the benefit of hearing aids* [Public sector] |
|  | *Understanding their ability to discriminate speech. Assessing abilities with HAs* [Public sector] |
|  | *For counselling and realistic expectations* [Public sector] |
|  | *Once performing verification, we can then validate the HA fitting through speech testing. This gives us valuable information as to how the HA is performing to ensure the pt is getting the right HA but we can also use it as a counselling tool.* [Public sector] |
|  | *Used to show outcome for bone conduction hearing aids* [Public sector] |
|  | *improved ability to counsel and adjust* [Public sector] |
|  | *Processing speech is usually the most important factor in our patient's hearing.* [Public sector] |
|  | *Assessment of functional impact of loss and the family notional benefit of hearing instruments.* [Private sector] |
|  | *A tangible test/result the client understands. It assesses the key/primary complaint of most clients.* [Private sector] |
|  | *I find it's a good counselling tool .* [Public sector] |
|  | *To let patients and their families to see the difficulties a patient may be having understanding speech* [Private sector] |
|  | *Patients/significant others seeing difficulties with hearing loss and benefit of hearing aids in more relatable way. Good counselling tool for clinicians.* [Public sector] |
|  | *The demonstrate to patient and significant other benefits of hearing aids vs. no hearing aid or 1vs.2.* [Public sector] |
|  | *To more quickly and objectively gauge whether a change in hearing aid/prescription has been useful* [Public sector] |
|  | *Confirming whether hearing aids are helping. Unaided testing under headphones can identify optimal speech recognition and be a useful counselling tool* [Public sector] |
|  | *If they are in denial it can be easier for them to see the results are correct* [Private sector] |
|  | *It can show the patient the improvement the hearing aids can make in regards to speech discrimination. It can also be a good gauge for the patient to find out how good or bad their hearing levels are. The speech test is given as a percentage score at the end of the test. This gives the patient some kind of numerical aspect to their results. An audiogram may not give them as much clarity, they may not fully understand it and displaying pure tones is far from complex sentences. It is a more simplified test which is easily understood and explained.* [Public sector] |
|  | *Invaluable for counselling and inform client of realistic expectations with hearing aid use.* [Private sector] |
|  | *Useful as a counselling tool when discussing realistic expectations of a hearing aid* [Public sector] |
|  | *More realistic + gives far more insight into how much potential benefit amplification may have for each individual* [Private sector] |
|  | *a really useful counselling tool with patients at assessment to explain their processing in noise without visual cues (especially those with a greater SNR loss). A shame it is not a verified test for assessing improvement with hearing aids.* [Private sector] |
|  | *Proving benefit to client and also ensuring that the best results have been achieved.* [Private sector] |
|  | *assesses real world difficulties* [Public sector] |
|  | *better knowledge of the benefit [*Private sector] |
|  | *It can help prove to the client the benefits of wearing hearing aids all the time* [Private sector] |
|  | *See the real hearing loss, may think others appreciate the real difficulties* [Public sector] |
|  | *Allowing Patient to see benefit of aids and also to help with rehab* [Private sector] |
|  | *Use to counsel clients on realistic expectations of hearing aids* [Private sector] |
|  | *Rehab/Counselling* [Public sector] |
|  | *Give patients realistic expectations.* [Public sector] |
|  | *To measure benefit to aiding and demonstrate it to the client* [Private sector] |
|  | *On the assessment stage - to get a more comprehensive understanding of the hearing difficulties and for the client to see what they are missing. On the fitting/follow up stage - to understand if the client is getting the desired benefit, for them to see the measured improvement and for better counselling.* [Private sector] |
|  | *pt understands the results and sees the results* [Private sector] |
|  | *speech recognition with and without aiding, checking clarity and sensitivity of aids checking pts processing capabilities.* [Public sector] |
|  | *Counselling ... show improvements when fitted with aids* [Private sector] |
|  | *patient feels that it is useful for hearing aid set up* [Public sector] |
|  | *can help counsel with realistic expectations* [Public sector] |
|  | *When i do speech testing it tends to be as a counselling tool to demonstrate to patients how the hearing aids are helping them. It can also help determine eligibility for cochlear implant criteria.* [Public sector] |
|  | *More realistic scenario of everyday listening compared to PTA. Reassurance of good performance in speech in noise. Highlight importance of good communication tactics such as lip reading and minimising background noise.* [Public sector] |
|  | *Gives you a better understanding of patients loss and possible problems, for both clinician, patient and family which should allow for better treatment outcome. Should reduce return appts for some patients and lead to better satisfaction for those often listed as difficult.* [Public sector] |
|  | *Diagnostic tool - pathologies* [Public sector] |
|  | *For rehabilitation, particularly severe/profound patients to help them understand the limitations for them* [Public sector] |
|  | *Gain a greater understanding of auditory pathway . Provide better counselling and manage expectations* [Private sector] |
|  | *There is a sometimes case for using speech testing as a tool for assisting the patient to more fully understand that they have a hearing loss that needs to be treated.* [Private sector] |
|  | *good counselling tool for both patient and significant other* [Public sector] |
|  | *For patients, it highlights need and reaffirms their awareness of hearing loss. For clinicians, it helps us understand their communication issues in a way a pure tone audiogram cannot. [Private sector]* |
|  | *could be used as a counselling tool or outcome scores* [Public sector] |
|  | *Good counselling tool and fine tuning of hearing aids* [Public sector] |
|  | *they are good as a counselling tool to show the benefit the patient is gaining, more as an assurance* [Public sector] |
|  | *As a check of real life hearing.* [Public sector] |
|  | *Evaluating speech discrimination and in turn benefit from hearing aids with patients that have sudden losses. Also a good counselling tool for patients.* [Public sector] |
|  | *counselling tool, measuring 'real-life' impact of changes to hearing aid fitting, quantifying 'real-world' difficulty that cannot be assessed from PTA (particularly for mild losses)* [Public sector] |
|  | *Being able to actually show the patients the results can be a really good tool - both positive and negative* [Public sector] |
|  | *I used to perform quick sin on every assessment, however going forward real-world testing in the client's own environment is much better. Sadly most clinics expect you to pay upfront for your aids. Our clients only pay if/when they are fully happy. Results of speech tests are useful for us as professionals, but I believe what the client thinks is a better gauge. [Private* sector] |
|  | *Counselling patient about their hearing loss, demonstrating benefit from hearing aids* [Public sector] |
|  | *A better representation of real world difficulties* [Public sector] |
|  | *show patients they are benefitting from hearing aids* [Public sector] |
|  | *Ensuring benefits of hearing aids* [Public sector] |
|  | *Counselling in relation to limitations of hearing aids. Information to support Cochlear Implant referral Information to support additional fine tuning in particular frequency lowering* [Public sector] |
|  | *Live speech is beneficial, we also take patients to an area with more noise to check speech in noise in real life setting* [Public sector] |
|  | *Verification of levels and shows pt and family difference made with aids* [Public sector] |
|  | *Counselling. Visual representation of limitations/hopeful improvement.* [Public sector] |
|  | *Validating fittings, counselling, checking eligibility for implants, evaluating performance in noise* [Public sector] |
|  | *better understanding of patients speech perception for the clinician, Counselling tool for pt + significant other present* [Private sector] |
|  | *Demonstrate the effectiveness of the hearing aids - useful as a counselling tool for patients* [Public sector] |
|  | *We aim to enable patients to hear speech the best they can with their residual hearing and invariably speech with be within noisy, challenging situations.* [Public sector] |
|  | *Counselling + proving benefit of intervention (relative to unaided condition)* [Public sector] |
|  | *demonstration of HA benefit* [Public sector] |
|  | *Counselling tool in that it relates more to their everyday experiences than PTA.* [Public sector] |
|  | *Multiple from a counselling perspective but due to multi-lingual issues we wouldn't be able to test our diverse range of patients so the true benefits wouldn't be realized for all users* [Public sector] |
|  | *Explain limitations to patients and their family who regularly attend complaining of lack of clarity if it is due to discrimination, not their hearing aids.* [Public sector] |
|  | *Demonstration to patients, family and carers of the benefit of hearing aids. Very useful to verify a variable PTA* [Public sector] |
|  | *I will always use speech testing for complex patients at the assessment and the fitting stage. The results provide a tool for me to assess potential for benefit from aid fitting, and also to detect any underlying reasons for poor benefit from aids or for patients describing hearing far worse than their PTA suggests. Speech testing is also useful for non-organic patients. At fitting stage (and follow up), I use BKB sentences with and without lipreading, aided and unaided. I find this incredibly helpful for guiding optimum fitting, and it also gives patients and their family/carers a really good indication of how much they use lipreading, how much the hearing aid is benefitting them, and this often leads into counselling on communication tactics as well.* [Public sector] |
|  | *Determine benefit gained In hearing aid amplification, counselling tool.* [Public sector] |
|  | *An additional set of results to inform hearing aid settings, general functional ability, and provide a more holistic assessment of patient needs.* [Public sector] |
|  | *For routine adults mostly counselling benefit - can demonstrate the positive impact of hearing aids to patients and family members or demonstrate hearing deficiencies in a more relatable way. Often used for diagnostics and cochlear implant assessments.* [Public sector] |
|  | *demonstrate benefit of aids - demonstrate limitations of aids* [Public sector] |
|  | *Provide objective results in how much a person could hear in BGN, and provides results in the types of words they find difficult to hear.* [Public sector] |
|  | *Objective way of assessing patients processing ability as well as showing the benefits of differing hearing aids* [Private sector] |
|  | *Functional assessment of the fitting and a great counselling tool* [Public sector] |
|  | *verification of the fitting on the day* [Public sector] |
|  | *counsel on realistic expectations of hearing aid(s). Validation of hearing aid benefit/improvement Establishing non -organic hearing loss Evaluate function of direction microphones on hearing aids Diagnoses of ear related pathology* [Public sector] |
|  | *It would be another tool to validate the fitting* [Public sector] |
|  | *sadly we do not perform speech unless patient is booked into SVP clinic. Speech testing would give more insight to SNR and SRT of the patient. It could also be used as a counselling tool for SII in quiet and in noise for aided/ unaided verification. Speech testing could be used for validation in aided situations* [Public sector] |
|  | *Being able to know which sounds patients struggle with and how often patients are missing speech, impacts fitting* [Public sector] |
|  | *Easier counselling tool* [Public sector] |
|  | *Helps to determine the benefits of hearing aids. Can use it to counsel the patients.* [Public sector] |
|  | *Useful information on need for directional microphones and lipreading ability of the patient. Shows significant other the difficulties experienced by the patient* [Public sector] |
|  | *Detection of non-organic hearing losses, retro cochlear hearing loss and understanding and explaining the benefits of aiding with patients with poor speech discrimination in quiet. I have also detected poor speech discrimination with a 6-8 KHZ which showed extremely poor speech discrimination on an ENT clinic. The patients speech discrimination improved after steriod treatments over a course of a few weeks* [Public sector] |
|  | *Real world concerns for patients is how well they hear conversation. Speech testing aided and unaided provides baselines to build a person centred specific rehab programme* [Private sector] |
|  | *Critical component to see how the hearing aids are working for that patient. Couldn’t work without them* [Private sector] |
|  | *Allows to a better understanding of the patient's daily performance and everyday challenges* [Private sector] |
|  | *Real life testing. Discrimination test far better than PTA where it is only detection* [Public sector] |
|  | *Counselling tool, measure of functional benefit* [Public sector] |
|  | *It is a good reflective tool. Can explain the improvement of the hearing accurately to the patient.* [Public sector] |
|  | *Helps understand difficulties in real life situations and gives and idea as what hearing aid and features would benefit the patient the most and make the best recommendation.* [Private sector] |
|  | *Oh my word, invaluable for patient and SO. Assessing challenges and managing expectations* [Private sector] |
|  | *Enormous - should be far more routine but something has to give with the amount of patients we are required to see. IT should be routine to escalate complex patients into speech audiometry.* [Public sector] |
|  | *Clearer idea of hearing loss. Showing potential for improvement with hearing aids Helping to explain the issues with hearing* [Private sector] |
|  | *I use it as a counselling tool to set expectations. I will also use AB wordlist testing on patients who have hearing aids but report no benefit to guide me in deciding whether it is worth continuing with hearing aid on that ear or if other avenues should be explored* [Public sector] |
|  | *Verifies how well the patient can hear speech and give an idea of also how the patient is hearing speech in background noise* [Public sector] |
|  | *Allows to access individual patient difficulties in understanding speech. This will allow audiologist to fit hearing aid appropriately.* [Public sector] |
|  | *It can be a useful tool for assessing processing difficulties and for counselling* [Public sector] |
|  | *rehab; also it can have face validity for pt when dealing with diagnostic or rehab questions; address certain diagnostic questions* [Public sector] |
|  | *Counselling, building better picture of difficulties for those adults with hearing difficulties but normal thresholds, measuring improvement from hearing aids objectively* [Public sector] |
|  | *Greater counselling method- can be sued to determine expectations and also validity of settings and options to improve SNR* [Public sector] |
|  | *Knowing the speech threshold pre-fitting is important for counsel the patient in terms on how much benefit he can expect from the hearing aids* [Public sector] |
|  | *Managing expectations, picking the correct technology/accessories if applicable* [Private sector] |
|  | *Better understanding of loss for patient and support* [Private sector] |
|  | *Speech testing is a good tool to measure the benefit from the hearing aid. If speech testing is carried out on the first fit appointment, we can adjust the hearing aid characteristics based on this and address some issues then and there which will improve patient satisfaction and can reduce follow up visits. Also speech testing gives you an idea about the hearing aid wearer’s cognitive ability to understand speech and hence is a good tool in counselling family members regarding the realistic expectations. To conclude, the ultimate aim of amplification is to improve the communication and thereby improve the quality of life and having speech testing as an outcome measure is so important for each hearing aid wearers to ensure maximum speech clarity and benefit out of hearing aids.* [Public sector] |
|  | *improved expectations* [Public sector] |
|  | *It gives the prognostic indication & outcome of hearing aids benefit.* [Public sector] |
|  | *Better counselling and tech options leads to realistic expectations, therefore better outcomes and more informed decision making. Greater face validity that I’m addressing actual complaint* [Public sector] |
|  | *Understanding their hearing loss, understanding their limitations, understanding the benefits of their hearing aid, realistic expectations..Also beneficial to the audiologist in giving rehab advice* [Private sector] |
|  | *Verification that amplification is right* [Public sector] |
|  | *Gives a more accurate sense of how the hearing loss effects their life* [Private sector] |
|  | *Counselling* [Public sector] |

| **Guiding the fitting of hearing aid technology and assistive devices** | *Aided vs unaided - test benefit with Has* [Public sector] |
| --- | --- |
|  | *Reflect real life, more than pta. Helps reassure aiding benefit. [Private* sector] |
|  | *Real life stimulus is often easier for the patient to relate to and also gives a better idea of the actual benefit of hearing aids to the patient. Also gives some idea of processing as opposed to just detection of sound.* [Public sector] |
|  | *Difficult to say as we do not perform them but wish we did. I think we would find less people with hearing aids returning for clarity* [Private sector] |
|  | *To ascertain the improvement of clarity when fitted .* [Private sector] |
|  | *To help select a suitable level of performance in a hearing aid and set realistic expectations for what this patient can achieve from a new hearing aid and the need to consider an FM system or other accessory* [Private sector] |
|  | *Help with wireless accessories Options and discussion with overall outcome with hearing aids* [Private sector] |
|  | *Optimising the hearing aids Rehab tool* [Public sector] |
|  | *they generally know how the instruments have helped or not.* [Public sector] |
|  | *additional information on hearing aid benefit to aid rehab process and discuss expectations and needs for ALDS/streaming devices* [Public sector] |
|  | *Be interesting to see and record the speech testing outcomes before and after amplification* [Public sector] |
|  | *Hearing aid benefit assessment* [Public sector] |
|  | *To improve hearing aid fittings.* [Public sector] |
|  | *monitoring performance and evaluating HA and or CI benefit* [Public sector] |
|  | *To diagnose the difference between poor hearing instrument setup and poor speech discrimination* [Public sector] |
|  | *further evidence of the help of hearing aids* [Public sector] |
|  | *Testing benefits of hearing aids and testing functional hearing.* [Public sector] |
|  | *functional hearing aided vs unaided benefit verified demonstrate to pts* [Public sector] |
|  | *More clarity from hearing aids* [Public sector] |
|  | *I think it is absolutely essential as part of the basic battery in order to add information to hearing aid candidacy. It is just not a commonly used UK practice.* [Public sector] |
|  | *Very close to real life.* [Public sector] |
|  | *Better hearing aid outcomes* [Public sector] |
|  | *To provide an idea of how much benefit a hearing aid will be particularly for sloping high freq losses and to indicate if there are any processing problems* [Public sector] |
|  | *really good as an outcome measure but also for demonstrating the hearing loss to the patient or the benefit of the hearing aid* [Public sector] |
|  | *- validation of hearing aids - better able to see benefits* [Public sector] |
|  | *More real world situation than PTA. Helpful for evaluating outcome, showing patients benefits of hearing aids, establishing possible neural/processing element to hearing loss.* [Public sector] |
|  | *Realistic situations which affect them rather that just pure tones* [Public sector] |
|  | *It’s a form of validation. REM’s are important but they do not tell us if the patient is hearing speech well with the devices* [Public sector] |
|  | *To help with cochlear implant referral. To identify need for further/other rehabilitation* [Public sector] |
|  | *Gives a better idea of px speech understanding and if done before and after fitting, how the hearing aids are benefitted the px.*  [Private sector] |
|  | *A realistic test, comparable to the real life problems they face* [Public sector] |
|  | *Helps you give a better idea of how a pt would do with an aid but also can help with deciding if to aid* [Public sector] |
|  | *Validation of HA fitting. Assess functional performance with and w/o HA . Counselling tool for patient and/or significant other during assessment and/or at follow up. To reflect performance of real-world and review whether ALDs required or further adjustment of fitting required.* [Public sector] |
|  | *Patients feel test is more ‘real world’* [Public sector] |
|  | *Gives level of functional hearing difficulty perhaps not revealed by PTA alone. Allows before and after aiding comparison to validate efficacy of hearing aids.* [Public sector] |
|  | *Validation and benefit realisation of hearing aids, further the understanding of the impact of hearing loss.* [Public sector] |
|  | *More realistic assessment of hearing abilities and challenges especially when tested in noise and delivered through a multiple speaker array to test hearing aid performance.* [Public sector] |
|  | *Great benefit to get an understanding of processing of speech with the hearing aids in compared to not wearing them.* [Public sector] |
|  | *More realistic test materials as patients can relate more. Also good validation tool to demonstrate benefits of amplification to patients*[Public sector] |
|  | *Speech testing is very useful because it can help you to program aids very well and detects where patient is struggling with the speech clarity.* [Private sector] |
|  | *Verification of aid benefit* [Public sector] |
|  | *Huge. Real life relatable testing with benefits in counselling. A lot of patients request either speech testing or speech in noise testing but we are unable to accurately perform this aided. Unaided ear specific information can be gained from QuickSIN but this information has little relevance on programming.* [Public sector] |
|  | *Verification that amplification is right*[Public sector] |
|  | *To ascertain how well people can understand speech with and without hearing aids* [Public sector] |
|  | *Good 'objective' function of speech awareness unaided versus aided. Also good to compare aids/settings Patient can see how well he/she has/has not performed before and after changes* [Public sector] |
|  | *excellent validation tools* [Private sector] |
|  | *Validation of hearing aid effectiveness* [Public sector] |
|  | *They can see the benefit of amplification rather than just using REM* [Public sector] |
|  | *How patient heard speech in contrast to how they hear puretone kn a regular hearing test* [Public sector] |
|  | **Pt's realistic expectations * If Pt wants only one H/A we can aid the best one *Less fine tuning appts* [Public sector] |
|  | *Showing S:N ratio loss for recommending accessory. Checking if there is useful/aidable hearing for deciding on mon/binaural. Client satisfaction* [Private sector] |
|  | *Verification of benefit. Monural or Binaural prescription* [Public sector] |
|  | *Extra information for hearing aid programming e.g if there is rollover - Extra information in guiding patients about assistive devices e.g remote mics* [Public sector] |
|  | *Gives some clues re real world expectations, informs CI referral decisions and informs advice re directional mics and FM.* [Public sector] |
|  | *It's a useful tool when explaining their hearing loss, expectations and limitations. It can reinforce the need for directional mics or fm systems* [Public sector] |
|  | *More real world validation of hearing aid fitting* [Public sector] |

| **Diagnostic Test Battery** | *More specific Audiological picture of patient's speech perception, also if carried out speech in noise testing then an idea of how patient manages in BGN compared to just a regular PTA* [Public sector] |
| --- | --- |
|  | *Be aware of underlying processing disorders* [Public sector] |
|  | *To assess weather there issues are solely due to threshold or if there are any other underlying issues impairing there hearing.* [Public sector] |
|  | *Ability to understand auditory processing ability which can help set expectation levels of both patient and clinician. You can also use these results to determine which hearing aid technology is required to solve the diagnosis best [Private sector]* |
|  | *Identifies patients with very poor speech discrimination abilities. Assists with counselling on cochlear implant candidacy.* [Public sector] |
|  | *In many ways a better litmus test than PTA for hearing impairment assessment. I intend to use it far more in the future as I see it as validating the benefits for patients of HAs - particularly in the early adaptation phase when challenges to long term continued use are most acute.* [Private sector] |
|  | *Diagnostic test for speech processing* [Private sector] |
|  | *-- identification of non organic hearing loss and processing disorders -* [Public sector] |
|  | *Rule out processing issues* [Public sector] |
|  | *Checking the processing of sounds and auditory pathway function* [Public sector] |
|  | *Useful to check speech discrimination pre surgery (Otosclerosis) or as counselling tool pre and post hearing aid fitting* [Public sector] |
|  | *Main use is to determine whether to refer for cochlear implants.* [Public sector] |
|  | *Can help to identify retrocochlear lesions 2. Can quantify the benefit of hearing aids* [Public sector] |
|  | *Looking at speech processing ability is very important for setting suitable expectations and choosing technology.* [Private sector] |
|  | *good tool for testing for Non organic, APD, and patients who report speech discrimination problems* [Public sector] |
|  | *Better understanding of how the individual ears and the binaural system uses complex sounds. I would not fit a hearing aid without testing speech initially.* [Public sector] |
|  | *To confirm type of loss, accuracy of results and help predict a more accurate reham program to follow as an individual* [Public sector] |
|  | *Use it for Adults with Learning Impairments - where PTA is not suitable* [Public sector] |
|  | *Identification of specific problems* [Public sector] |
|  | *The standardised PTA doesn't reflect realistic listening situations and so speech tests may provide a more accurate representation of an individual's hearing difficulties* [Private sector] |
|  | *Speech recognition and discrimination* [Public sector] |
|  | *It provides a functional element to diagnostic testing of hearing, and rehabilitation.* [Public sector] |
|  | *Better assessment of functional abilities.* [Public sector] |
|  | *Checking processing ability* [Public sector] |
|  | *More diagnostic information* [Public sector] |
|  | *appreciation of Pt processing ability apart from aided/hearing ability* [Public sector] |
|  | *Often patients main problem is understanding speech, especially in background noise, so being able to measure this as part of a standard appointment may be valuable e.g. to measure HA benefit* [Private sector] |
|  | *I think the Quick-SIN and the short-version LiSN - S with prescribed gain amplification are important and valuable measures of SNR-loss. These measures in my opinion provide a greater insight into patient difficulties. We should not be basing rehabilitation solely on the PTA.* [Private sector] |
|  | *We are sometimes asked by ENT to perform speech testing on acoustic neuroma clinics, but do not routinely use them with direct access patients unless there is a complex patient or unexplained problems with hearing , i.e to assess if there may be an auditory processing issue. Can help assess whether processing is an issue and so to explain this to patients.* [Public sector] |
|  | *Good rehabilitation tool - More ecological validity - Assess speech intelligibility and more suitable amplification choice - Assess hearing aid benefit [*Public sector] |
|  | *Patients with certain conditions like vestibular schwannoma, Meniere's disease or just a sudden hearing loss, may all encounter distortion so it is helpful to know whether a conventional aid is appropriate or a CROS instead and also for managing pt expectations/counselling tool.* [Public sector] |
|  | *Measurable outcome comparison, a functional test for speech discrimination* [Public sector] |
|  | *It's what we hear day to day, we do not hear puretones in real life. More of a measure of day to day sounds.* [Public sector] |
|  | *we use it to identify prospective middle ear surgery patients* [Public sector] |
|  | *Useful as part of a diagnostic test battery* [Public sector] |
|  | *To assess Hearing difficulty in noise To assess CI candidacy* [Public sector] |
|  | *It would be another tool to validate the fitting* [Public sector] |
|  | *sadly we do not perform speech unless patient is booked into SVP clinic. Speech testing would give more insight to SNR and SRT of the patient. It could also be used as a counselling tool for SII in quiet and in noise for aided/ unaided verification. Speech testing could be used for validation in aided situations* [Public sector] |
|  | *Identifying speech discrimination difficulty when normal PTA or benefit of hearing aid use (eg if dead regions, acoustic neuroma)* [Public sector] |
|  | *Good tool in conjunction with PTA for diagnosis and predicting hearing aid outcomes* [Public sector] |
|  | *allow you to see true hearing not just pta* [Public sector] |
|  | *can help to assess level of functional hearing, and the assessment of rollover for retrocochlear disorders* [Public sector] |
|  | *Further investigation of hearing. Pure tone audiometer is more like a reflex compared to listening>cognition>repetition. Doing speech tests help me to screen for complex cases and APD-type patients too.* [Private sector] |
|  | *Speech in noise testing gives a realistic perspective on your patients challenges* [Private sector] |
|  | *Identify rectrocochlear pathology if result doesn't match up to audiogram or identify malingerer* [Public sector] |
|  | *level of intelligibility poor discrimination with regressive understanding at high intensities* [Private sector] |
|  | *It’s the only way to understand the functional impact. It can be very surprising how much the speech test scores can vary from the audio gram in patients who are not successful with amplification.* [Public sector] |
|  | *Dealing with the real concerns of the patient. Genuine understanding of patient needs and what technology may or may not do for them.* [Public sector] |
|  | *Diagnostics, part of APD test battery, aural rehabilitation (cognitive load and Listening effort, directional microphones, noise reduction algorithms, validation of fitting etc)* [Public sector] |
|  | *They are able to correlate based on their own experiences (challenging listening situations).* [Public sector] |
|  | *To demonstrated the benefit of the hearing aids in aided and unaided conditions* [Public sector] |
|  | *To assess overall hearing function and APD* [Public sector] |
|  | *Detection of cochlear damage/dysfunction, for hearing aid amplification outcome, with differential counselling if poor speech discrimination, and referral for CI assessments. Directional microphone settings.* [Public sector] |
|  | *Pure tone audiogram is not a sufficient indicators of speech perception ability. Without testing speech we have not addressed most patients primary complaint. We cannot say we have fully assessed the patient or provide appropriate counselling without doing speech testing* [Public sector] |

Alanazi, A.A. (2017). Audiology and speech-language pathology practice in Saudi Arabia. *Int J Health Sci (Qassim), 11*(5), 43-55.

Ali, A., Hickson, L., & Meyer, C. (2017). Audiological management of adults with hearing impairment in Malaysia. *International Journal of Audiology, 56*(6), 408-416. doi:10.1080/14992027.2017.1305515

American Speech-Language-Hearing Association. (2019). Audiology survey report: private practice. Retrieved from <https://www.asha.org/siteassets/surveys/>

Anderson, M.C., Arehart, K.H., & Souza, P.E. (2018). Survey of Current Practice in the Fitting and Fine-Tuning of Common Signal-Processing Features in Hearing Aids for Adults. *J Am Acad Audiol, 29*(2), 118-124. doi:10.3766/jaaa.16107

DeBow, A., & Green, W. (2000). A Survey of Canadian Audiological Practices : Pure Tone and Speech Audiometry. *Canadian Journal of Speech-Language Pathology and Audiology, 24*(4), 153-161.

Easwar, V., Boothalingam, S., Chundu, S., Manchaiah, V., & Ismail, S. (2013). Audiological Practice in India: An Internet-Based Survey of Audiologists. *Indian Journal of Otolaryngology and Head & Neck Surgery, 65*. doi:10.1007/s12070-013-0674-2

Kirkwood, D.H. (2005). When it comes to hearing aids, “more” was the story in '04. *The Hearing Journal, 58*(5). Retrieved from <https://journals.lww.com/thehearingjournal/Fulltext/2005/05000/When_it_comes_to_hearing_aids,__more__was_the.5.aspx>

Martin, F., Champlin, C., & Chambers, J. (1998). Seventh survey of audiometric practices in the United States. *Journal of the American Academy of Audiology, 9*(2), 95.

Myles, A.J. (2017). The clinical use of Arthur Boothroyd (AB) word lists in Australia: exploring evidence-based practice. *International Journal of Audiology, 56*(11), 870-875. doi:10.1080/14992027.2017.1327123

Nandurkar, A., Mukundan, G., & Gore, G. (2015). Speech perception assessment practices among Audiologists in India: A Preliminary survey. *International Journal of Speech and Language Pathology and Audiology, 3*, 52-65.

Thakor, H. (2020). *South African Audiologists’ Use of Speech-in-Noise Testing for Adults with Hearing Difficulties.* (MSc Audiology). University of the Witwatersrand,
